# Healthcare resource utilization and use of biologics in chronic spontaneous urticaria

**DOI:** 10.1101/2025.06.06.25329137

**Authors:** Lillian D. Sun, Ajay Behl, Peter N. Danilov, Fatai Y. Agiri, Kathryn M. Pridgen, Amanda Christine F. Esquivel, Julie A. Lynch, Elma Baron

## Abstract

**Importance:** Biologic therapies have emerged as a promising treatment for chronic spontaneous urticaria (CSU), a debilitating skin disease. However, real-world data on their use is limited.

**Objective:** Analyze the time from diagnosis to biologic therapy initiation and the CSU-specific healthcare resource utilization (HCRU)

**Design:** Retrospective cohort study

**Setting:** Veterans Health Administration (VHA) from January 2011 through December 2021

**Participants:** Veterans ≥18 years of age with ≥2 urticaria diagnoses or one urticaria and one angioedema diagnosis with no diagnosis of urticarial vasculitis

**Exposures:** Diagnosis of CSU, defined as ≥2 urticaria diagnoses (ICD9 codes: 708.1, 708.8, 708.9; ICD10 codes: L50.1, L50.8, L50.9) or one urticaria and one angioedema (ICD9 code: 995.1; ICD10 code: T78.3) diagnosis

**Main outcomes and measures:** Time from diagnosis to biologic initiation and CSU-specific outpatient visits, inpatient admissions, emergency room visits, and pharmacy claims, in the 12-month pre- and post-index periods

**Results:** The final cohort included 26,387 Veterans with CSU, with a mean age of 54.9 years (SD=15.2). In the 12 month post-index period, 23,699 Veterans (89.8%) started treatment, but only 613 Veterans (2.6%) started biologic therapy, with a median initiation time of 337 days. CSU-specific HCRU increased notably in the post-index period across all categories. 66.8% of Veterans had pharmacy claims pre-index date compared to 89.8% post-index date. Outpatient visits were utilized by 92.4% of Veterans pre-index date and 96.7% post-index date.

**Conclusion and relevance:** The findings suggest that CSU management may be further optimized within the first year of diagnosis and that initiation of biologics may be considered sooner in appropriate patients. Additionally, the increased HCRU observed in the post-index period highlights the burden that CSU places on both patients and the healthcare system.

## Introduction

Chronic spontaneous urticaria (CSU) is a debilitating skin disease characterized by recurrent wheals, angioedema, or both for six weeks or longer without an identifiable external cause. The clinical presentation includes persistent itching and pain, sometimes leading to disrupted sleep and chronic fatigue.^1^ This constellation of symptoms with unpredictable flare-ups and lack of a clear cause significantly impacts patients daily, affecting work, social activities, and emotional well-being. The psychological burden of CSU is profound, leading some to social withdrawal and an overall decreased quality of life.^1–3^

Although biologic therapies have emerged as a promising treatment for moderate to severe CSU, particularly in patients unresponsive to conventional therapies,^4^ real-world data on their use is limited. This study addresses this gap by analyzing the time to biologic therapy initiation and healthcare resource utilization (HCRU) in Veterans with CSU receiving care through the Veterans Health Administration (VHA).

## Methods

### Data Source and Ethics Statement

This retrospective cohort study leveraged data from the Corporate Data Warehouse (CDW), which provides access to longitudinal electronic health record data from the VHA, the largest integrated healthcare system in the United States.^5,6^ This study received ethical approval from the Minneapolis and Salt Lake City VA Institutional Review Boards. All patient information was de-identified and patient consent was not required.

### Study population

This study uses data from January 1, 2011, to December 31, 2021 and utilized an established algorithm to identify CSU cases.^7^ Participants were required to be ≥18 year and have ≥2 urticaria diagnoses (ICD9 codes: 708.1, 708.8, 708.9; ICD10 codes: L50.1, L50.8, L50.9) or one urticaria and one angioedema (ICD9 code: 995.1; ICD10 code: T78.3) diagnosis. Diagnoses must have been made ≥6 weeks but ≤12 months apart. The Veterans were also required to have ≥12 months of continuous care before and after the index date, defined as the first date with an associated CSU diagnosis. Patients with urticarial vasculitis (ICD10 code: L95) or who were missing date of birth or gender information were excluded.

### Data Collection and Analysis

We retrieved patient demographics at index date and clinical characteristics of patients, including treatments received during the 12 month pre- and post-index periods. The cohort was analyzed for time to biologic initiation and CSU-specific HCRU, measured as CSU-specific outpatient visits, inpatient admissions, emergency room visits, and pharmacy claims, in the 12-month pre- and post-index periods. Data were quantified using descriptive statistics.

## Results

### Demographics and Prior Treatment Use

We identified 137,749 patients with a diagnosis of CSU during the study period. After applying inclusion criteria, the cohort included 26,597 Veterans. We excluded those with a primary diagnosis for urticarial vasculitis at any time during the study period (n=208) and those with a missing value for patient ID, year of birth, or gender (n=2), resulting in a final cohort of 26,387 patients with a mean age of 54.9 years (SD=15.2). Most patients were male (76.4%), though CSU was more common in women (0.37%) than men (0.18%). Among the cohort, 60.8% (n= 16,048) self-identified as White and 28.0% (n= 7395) as Black. (Table 1)

**Table 1.** Veteran Demographic Data.

| **Characteristics** | **Veterans with CSU**<br>**(N=26,387)** |
| --- | --- |
| **Age, mean (SD), years** | 54.9 (15.2) |
| **Age category, n (%)** |  |
| 18-29 years | 1473 (5.6) |
| 30-39 years | 3913 (14.8) |
| 40-49 years | 4399 (16.7) |
| 50-59 years | 5557 (21.1) |
| 60-69 years | 6868 (26.0) |
| ≥70 years | 4177 (15.8) |
| **Male, n (%)** | 20,149 (76.4) |
| **Self-reported race, n (%)** |  |
| White | 16,048 (60.8) |
| Black | 7395 (28.0) |
| Asian | 723 (2.7) |
| Native Hawaiian/Pacific Islander | 324 (1.2) |
| American Indian/Alaska Native | 196 (0.7) |
| **BMI, mean (SD), kg/m<sup>2</sup>** | 30.2 (6.3) |
| **BMI category, n (%)** |  |
| Underweight: <18.5 kg/m<sup>2</sup> | 336 (1.3) |
| Normal weight: 18.5 to <25 kg/m<sup>2</sup> | 4731 (17.9) |
| Overweight: 25 to <30 kg/m<sup>2</sup> | 8793 (33.3) |
| Obese: ≥30 kg/m<sup>2</sup> | 12,229 (46.3) |
| **Smoking status, n (%)** |  |
| Current smoker | 5566 (21.1) |
| Former smoker | 6920 (26.2) |
| Never smoker | 8961 (34.0) |

Most Veterans with CSU were from the southern US (46.5%), followed by the west (19.9%), with 71.9% living in urban areas. Comorbidities were common; many were classified as overweight (33.3%, n=8793) or obese (46.3%, n=12,229), and 21.1% (n=5566) were current smokers, highlighting lifestyle risk factors.

In the 12 month post-index period, 23,699 Veterans (89.8%) started treatment. The most common treatments before and after the index date were antihistamines, corticosteroids, leukotriene receptor antagonists, tricyclic antidepressants, and immunosuppressants like cyclosporine (Figure 1). Only 613 Veterans (2.6%) started biologic therapy, with a median initiation time of 337 days.

**Figure 1.**
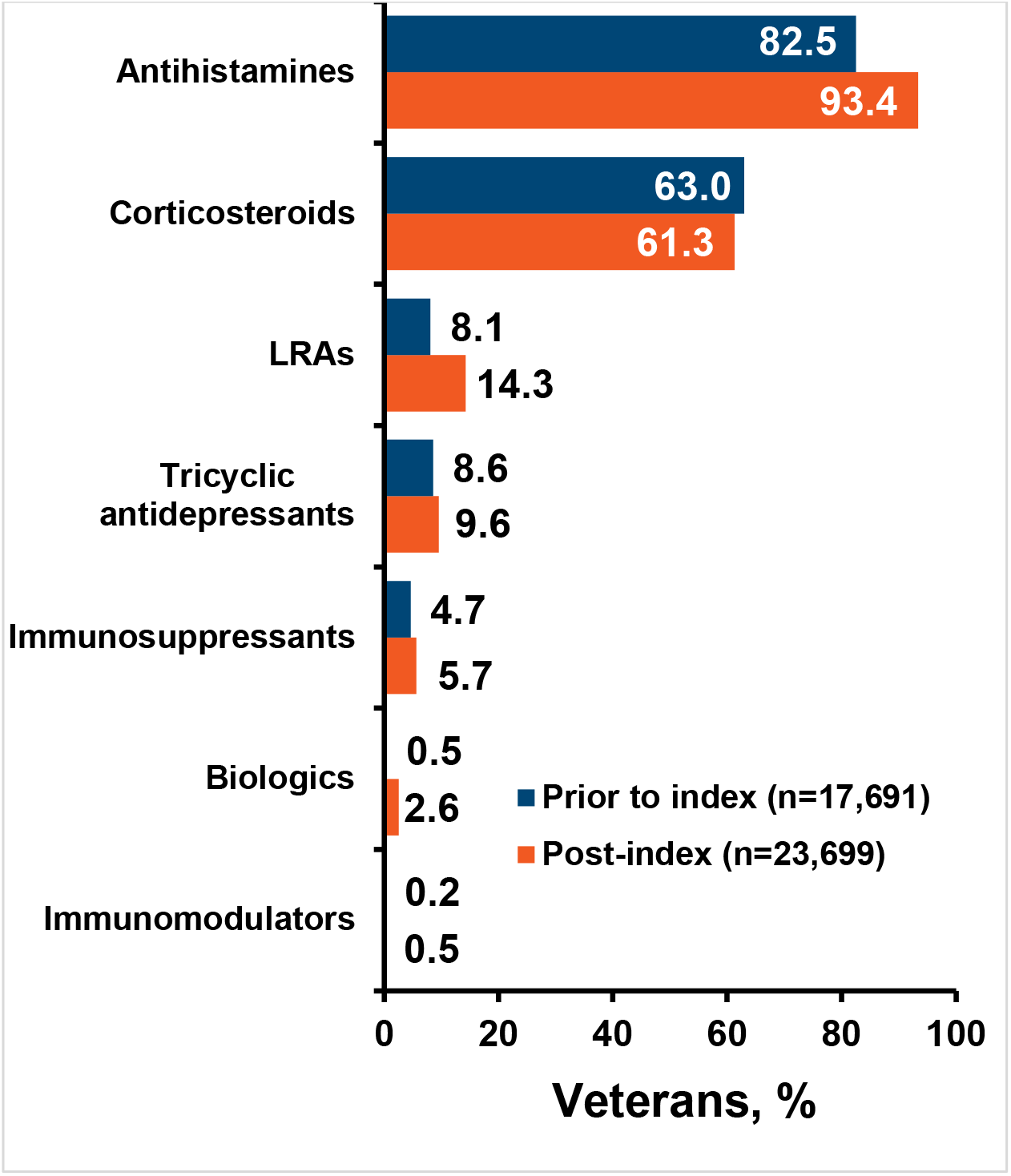
Pharmacological Treatments for Veterans with CSU

### Healthcare Resource Utilization (HCRU)

We measured HCRU by outpatient visits, inpatient admissions, ER visits, and pharmacy claims before and after CSU diagnosis. CSU-specific HCRU increased notably in the post-index period across all categories (Figure 2). 66.8% of Veterans had pharmacy claims pre-index date compared to 89.8% post-index date, indicating increased medication use. Outpatient visits were utilized by 92.4% of Veterans pre-index date and 96.7% post-index date, highlighting the need for ongoing care. Inpatient admissions and ER visits also rose, with the median length of stay increasing from four to five days, reflecting the potential severity of CSU episodes.

**Figure 2.**
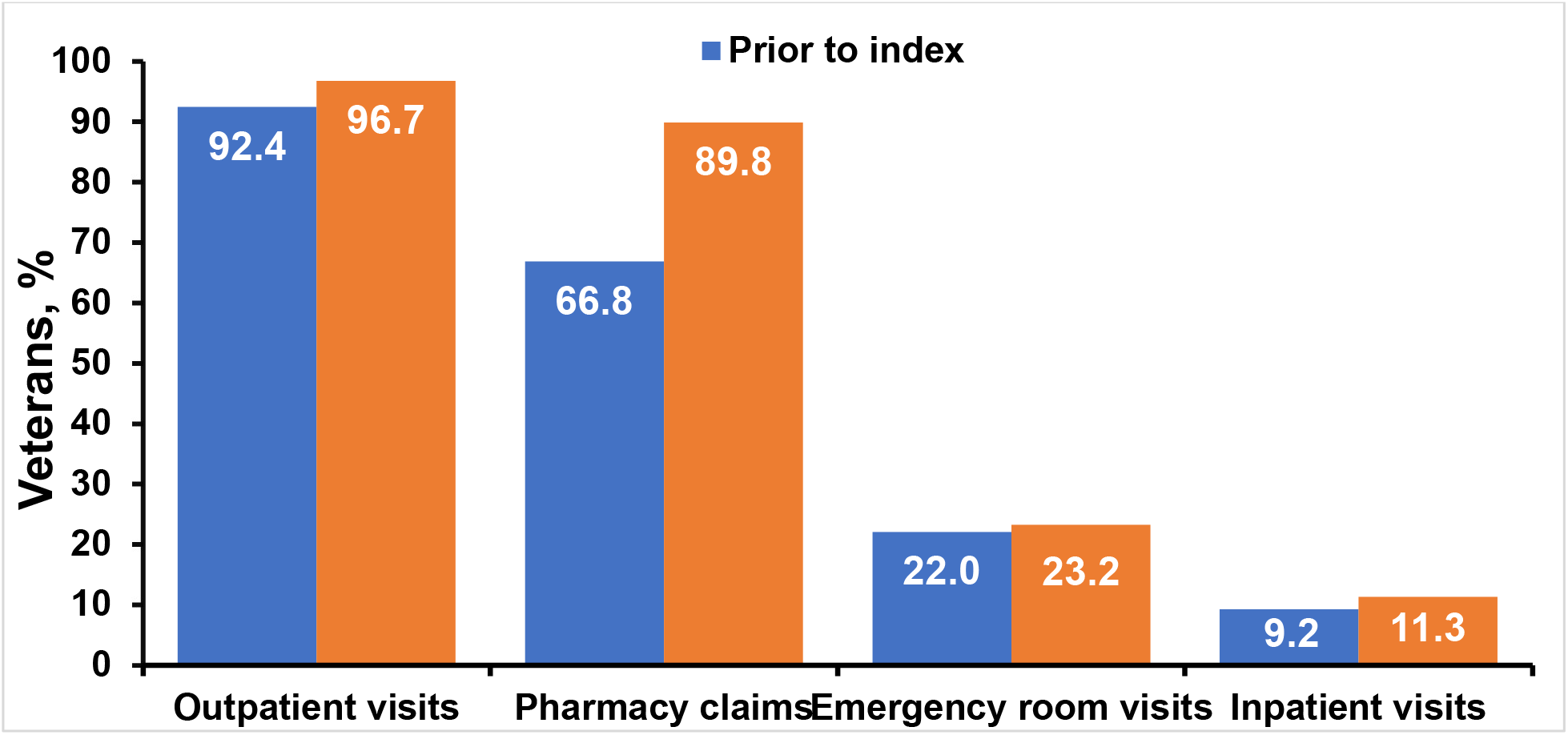
Healthcare Resource Utilization Among Veterans with CSU

## Discussion

The results of this study highlight two key findings: first, the potential underutilization of biologic therapies among US Veterans with CSU and second, the significant healthcare resource burden associated with CSU management.

The low percentage (2.6% within 12 months) of patients with CSU receiving biologics suggests a continued reliance on other treatments, such as antihistamines and corticosteroids, despite the fact that antihistamines have been observed to be insufficient for a significant portion of the population and corticosteroids likely increase adverse events.^8–10^ While potentially useful in managing some symptoms of CSU, these drugs may fail to provide long-term disease control for patients with moderate to severe CSU who need more targeted therapeutic strategies.^11^ This delay could be due to various factors, including patient hesitancy to utilize injectable medications, limited awareness of newer treatment options, or level of comfort on the part of the provider, which may greatly differ among primary care providers compared to dermatologists and allergologists.

Similar to previous research indicating a higher healthcare burden among patients with CSU,^1^ our study found increased HCRU following diagnosis, underscoring the significant burden that CSU places on the healthcare system. The rise in pharmacy claims and outpatient visits indicate that a subset of patients may experience persistent symptoms due to suboptimal disease control. Optimizing treatment through more targeted therapies could potentially result in better disease control, thus reducing the overall demand on healthcare resources.

### Limitations

Some Veterans may have sought care outside the VHA or moved between facilities, which could cause inaccuracies. Additionally, VHA data is sourced from multiple platforms, potentially leading to incomplete information. This Veteran patient population also differs from the general public in age, comorbidities, and socioeconomic factors, affecting generalizability. Similarly affecting generalizability, the Veteran population is primarily male, but CSU is more common among women.^12^

## Conclusion

This study offers valuable insights into real-world treatment patterns and healthcare resource utilization of patients with CSU. The findings suggest that management of CSU may be further optimized within the first year of diagnosis and that initiation of biologics may be considered sooner in the appropriate patient. Additionally, the increased HCRU observed in the post-index period highlights the burden that CSU places on both patients and the healthcare system. Further research is needed to understand the barriers to biologic therapy initiation in both primary care and specialty clinic settings and to develop strategies to improve access to advanced treatment options for CSU. Optimizing treatment pathways for patients with CSU has the potential to improve patient outcomes and reduce the long-term healthcare resource burden associated with this chronic condition.

## Data Availability

Patient-level data are already accessibly to all VA researchers with appropriate IRB approvals.

## Notes

**Funding sources:** This work was supported by Novartis Pharmaceuticals Corporation.

**Conflicts of interest:** JAL, PND, FYA, and KMP report grants from Alnylam Pharmaceuticals, Inc., AstraZeneca Pharmaceuticals LP, Biodesix, Inc, Janssen Pharmaceuticals, Inc., Novartis International AG, Parexel International Corporation through the University of Utah or Western Institute for Veteran Research outside the submitted work.

### Competing Interest Statement

JAL, PND, FYA, and KMP report grants from Alnylam Pharmaceuticals, Inc., AstraZeneca Pharmaceuticals LP, Biodesix, Inc, Janssen Pharmaceuticals, Inc., Novartis International AG, Parexel International Corporation through the University of Utah or Western Institute for Veteran Research outside the submitted work.

### Funding Statement

This work was supported by Novartis Pharmaceuticals Corporation.

### Author Declarations

This study received ethical approval from the Minneapolis and Salt Lake City VA Institutional Review Boards. All patient information was de-identified and patient consent was not required.

